# Investigating the relationship between interventions, contact patterns, and SARS-CoV-2 transmissibility

**DOI:** 10.1101/2021.11.03.21265876

**Authors:** Filippo Trentini, Adriana Manna, Nicoletta Balbo, Valentina Marziano, Giorgio Guzzetta, Stefano Merler, Marco Ajelli, Piero Poletti, Alessia Melegaro

## Abstract

**Background:** After a rapid upsurge of COVID-19 cases in Italy during the fall of 2020, the government introduced a three-tiered restriction system aimed at increasing physical distancing. The Ministry of Health, after periodic epidemiological risk assessments, assigned a tier to each of the 21 Italian regions and autonomous provinces (AP). It is still unclear to what extent these different measures altered mixing patterns and how quickly the population adapted their social interactions to continuous changes in restrictions.

**Methods and findings:** We conducted a survey between July 2020 and March 2021 to monitor changes in social contact patterns among individuals in the metropolitan city of Milan, Italy, which was hardly hit by the second wave of COVID-19 pandemic. The number of contacts during periods characterized by different levels of restrictions was analyzed through negative binomial regression models and age-specific contact matrices were estimated under the different tiers. Relying on the empirically estimated mixing patterns, we quantified relative changes in SARS-CoV-2 transmission potential associated with the different tiers.

As tighter restrictions were implemented during the fall of 2020, a progressive reduction in the mean number of contacts recorded by study participants was observed: from 16.4% under mild restrictions (yellow tier), to 45.6% under strong restrictions (red tier). Higher restrictions levels were also found to increase the relative contribution of contacts occurring within the household. The SARS-CoV-2 reproduction number was estimated to decrease by 18.7% (95%CI: 4.6-30.8), 33.4% (95%CI: 22.7-43.2), and 50.2% (95%CI: 40.9-57.7) under the yellow, orange, and red tiers, respectively.

**Conclusions:** Our results give an important quantification of the expected contribution of different restriction levels in shaping social contacts and decreasing the transmission potential of SARS-CoV-2. These estimates can find an operational use in anticipating the effect that the implementation of these tiered restriction can have on SARS-CoV-2 reproduction number under an evolving epidemiological situation.

## INTRODUCTION

During the COVID-19 pandemic, one of the main strategies used by governments to reduce SARS-CoV-2 transmission has been the introduction of non-pharmaceutical interventions to favor physical distancing. When a second wave of COVID-19 started spreading in Italy in the fall of 2020, the government progressively enhanced measures aimed at increasing physical distancing [1,2,3,4]. To better respond to a geographically heterogeneous increase of the number of COVID-19 cases, a three-tiered restriction system was introduced on November 6, 2020. Since then, every week, a tier was assigned to each of the 21 Italian regions and autonomous provinces (AP) by the Ministry of Health after an epidemiological risk assessment based on a combination of quantitative indicators, such as the estimated level of transmission and the burden on the healthcare system [4]. The sets of measures adopted in the three tiers were labeled according to a color scheme: yellow, orange, and red, corresponding to increasing levels of restrictions. These included the reinforcement of distance learning in primary and secondary schools, as well as the introduction of restrictions of individuals’ mobility ranging from a ban on inter-regional movements to a stay-home mandate for the entire day [5].

Several studies conducted during the COVID-19 pandemic have quantified mixing patterns under different non-pharmaceutical interventions, highlighting the dramatic decrease of individuals’ contact rates with respect to pre-pandemic levels [6,7,8,9,10,11,12,13,14,15,16]. However, it is not well quantified to what extent different measures promoting physical distancing alter mixing patterns and how quickly the population adapt their social interactions to continuous changes in restrictions. A fundamental question that is still open is whether the change of mixing patterns associated to implemented restrictions can be used to anticipate the effect of a given intervention on SARS-CoV-2 transmissibility.

The present study aims at investigating the changes in number of contacts and mixing patterns linked to the gradual tightening of non-pharmaceutical interventions adopted by the Italian government during the second wave of the COVID-19 pandemic, and to examine their impact on SARS-CoV-2 transmission. To this aim, we performed a contact survey in the metropolitan city of Milan, Italy, between July 2020 and March 2021. The survey collected information on the number and type of daily contacts made by study participants over the course of the pandemic when interventions of different intensity were in place. This data allowed us to assess the impact of the adopted restrictions on human mixing patterns and, through the use of mathematical modeling, to estimate their effect on SARS-CoV-2 net reproduction number.

## METHODS

The study was conducted between July 10, 2020 and March 31, 2021 in the metropolitan city of Milan, in Lombardy Region of Italy. Data were collected in collaboration with Centro Medico Sant’Agostino (CSM), a private healthcare center that is part of the National Health Service.

### Tiered restrictions

During the period of data collection, the study population underwent all the restrictions levels defined by the Italian tiered restrictions system (Table S1 in Appendix). In the baseline analysis, we assimilated the pre-tier period between October 26 and November 5, 2020 to the yellow tier, due to the similar restrictions preventively introduced by the Lombardy region with a decree of October 16, 2020 [17]. In Appendix, we present the results obtained by considering this period separately and labeling it with a different color: green. The period between July 10, 2020, and October 25, 2020, is hereafter denoted as white tier.

### Study design and data collection

Participants were selected among individuals who booked an appointment to undergo voluntary IgG serological COVID-19 testing at CSM. While booking the serological test, individuals were invited to participate in the study and, after acceptance, to fill in an online questionnaire on their social behavior (see Appendix). Informed consent was sought for individuals aged 18 years or more, and from a parent or legal guardian for underage individuals. Only 696 (39%) of individuals who accepted to participate in the contact study eventually underwent the serological test, among those 462 (66%) received the result before filling in the contact questionnaire.

The questionnaire was composed of two parts: (i) in the first, we collected key socio-demographic information on the participant (age, gender, occupational status, household size, and age of their household members); (ii) in the second, we recorded information on contacts that the participant experienced during the day preceding the questionnaire. The questionnaire was implemented through an online platform. Data were anonymized by CSM before conducting the analysis of collected records.

To make our results comparable with pre-pandemic Italian data [18], we defined a contact as a physical interaction or a two-way conversation of at least five words in the physical presence of another person. For each reported contact, the following information was recorded: (i) the sex of the contacted person, (ii) the age (either precise or range) of the contacted person, (iii) if the contact happened indoor or outdoor, (iv) the frequency at which the contact usually happens (more than once a day, once a day, more than once a week, once a week, less than once a week), (v) if the experienced contact was physical or not, (vi) the relation between the study participant and the contacted person (household member, other relative, classmate, colleague, friend/partner, other), and (vii) the place of the contact (home, school, workplace, public transportations, or other).

Ethical approval for this study was waived by the Ethical Review Board of the Bocconi University, Milan.

### Analysis

#### Descriptive analysis of contacts

We analyze the frequency distribution of contacts of different participants for a set of covariates, including their age, sex, employment status, their serological status before the interview, and the restriction tier at time of interview. Repeated encounters reported with the same individual counted as one contact only, following the same approach used elsewhere [19, 20].

To assess differences across multiple groups, we use one-way ANOVA, followed by post-hoc Tukey test. Estimated 95% confidence intervals and p-values are based on the Studentized range statistic and the Tukey’s ‘Honest Significant Difference’ method.

We use negative binomial regressions to estimate the mean number of contacts as a function of the different covariates. Negative binomial regressions were preferred over Poisson regressions given evidence of overdispersion (variance > mean), and a significant likelihood ratio (P < 0.05) for the overdispersion parameter. Separate regressions were applied to the number of contacts reported overall, those occurred within and outside the participant household.

#### Contact patterns

We assigned a tier to each participant based on the one assigned to the Lombardy region at the time of conducting the interview. We computed the mean number of contacts reported by respondents after grouping by age (one age group for individuals between 0 and 20 years of age, five 10-year age groups from 20 to 69 years, and one for individuals aged 70 years or older) and by tier (white, yellow, orange, and red). Age-specific contact matrices were estimated considering both physical and non-physical contacts and adjusted for reciprocity following the approach used in [19,20]. Variability was explored by computing 1,000 bootstrapped contact matrices [7,20], where each bootstrap was obtained by sampling with replacement a number of interviews equal to the original sample size, choosing the age of the participant with probability proportional to the age distribution in the metropolitan city of Milan [20,7].

##### Estimation of reproduction number

The reproduction number associated with each tier was estimated by using the Next Generation Matrix approach applied to an age-structured SIR transmission model where interactions between individuals of different ages are defined by the contact matrices derived in this study [21]. A generation time of 6.6 days was assumed to reflect the serial interval of COVID-19 observed in the region at the beginning of the pandemic [22, 23]. The NGM was computed under the illustrative condition of a fully susceptible population and adjusted for heterogeneous susceptibility to infection at different ages, as estimated in [24]; homogeneous susceptibility at different ages was explored as a sensitivity analysis.

To evaluate relative changes in the transmission potential determined by the different tiers, we considered a baseline NGM, computed by using contact records collected before the introduction of tighter restriction in the Lombardy region (between July 13, 2020, and October 25, 2020). The reduction in transmission led by mild, moderate, and strong restrictions (yellow, orange, and red tiers, respectively) was then defined as one minus the ratio between the dominant eigenvalue of the NGM computed using contact patterns estimated for the considered tier and the one associated with the baseline NGM.

## RESULTS

### Description of study participants

A total of 1,785 individuals were interviewed. Due to the constrained nature of the study, female individuals (57.8%) and adults between 20 and 64 years of age (91.0%) were oversampled, as shown in Figure S1 in Appendix. The median age of participants was 37 years (IQR 30-48). Participants under 20 years of age and above 64 years of age represent the 2.7% (49 participants) and the 6.3% (112 participants) of sample, respectively.

Overall, 1,063 participants (59.6%) were interviewed before the introduction of the tiered restrictions system (white tier), 314 while in the yellow tier (17.6%), 178 while in the orange tier (10.0%) and 230 while in the red tier (12. 9%).

Most participants were employed (77.4%), 10.1% were students, and 12.5% either inactive or unemployed; most study participants (57.9%) were cohabiting with one individual. 40 study participants (2.2%) reported to have tested positive for SARS-CoV-2 IgG antibodies prior to the interview. Descriptive statistics are reported in Table 1.

**Table 1.**
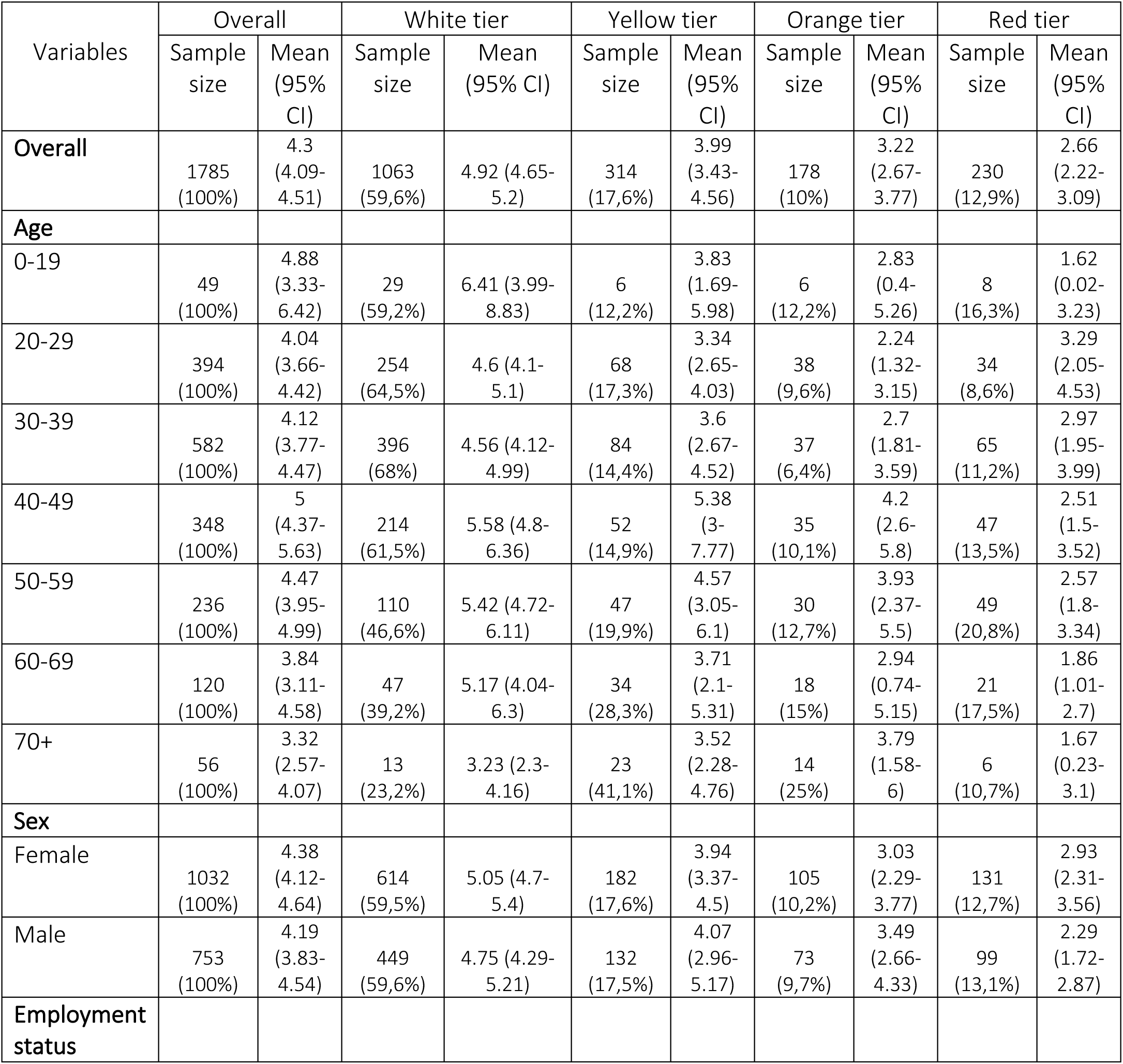
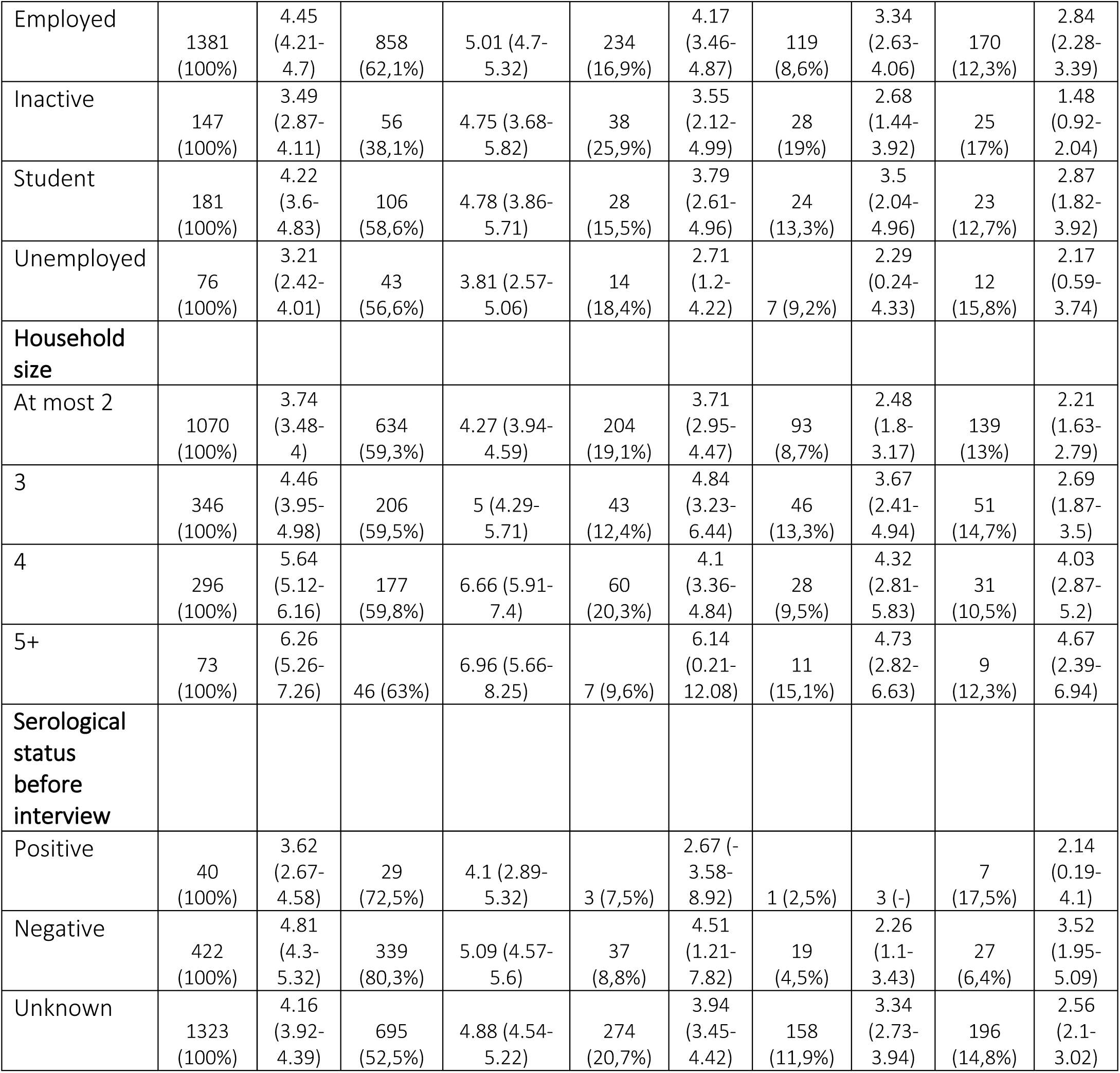
Sample description and mean number of daily contacts per person as recorded under different tiers.

### Contacts

The introduction of the tiered restriction system strongly influenced the number of social interactions of study participants of all age classes (see Figure 1A and Table 1). The mean number of contacts reported in white tier was 4.92 (95%CI: 4.65-5.20). The same quantity decreased by 0.93 (95% CI: 0.19-1.67), 1.70 (95%CI: 0.77-2.64) and 2.27 (95%CI: 1.43-3.11) during the yellow, orange, and red tiers, respectively (p-values for a post-hoc Tukey test applied to compare different means <=0.001). Compared to contacts observed before the introduction of the tiered restriction system, this corresponds to a relative reduction in the overall number of contacts of 18.9%, 34.6% and 45.9% for mild, moderate, and strong restrictions, respectively. Most marked changes were observed among individuals younger than 20 years, with a mean number of contacts decreasing from 6.41 (95%CI: 3.99-8.83) while in the white tier to 1.62 (95%CI: 0.02-3.23) while in the red tier. Individuals aged 20-29 years resulted those who were least affected by the tier change. Inactive individuals showed a higher reduction of contacts than students and employed individuals (68.8% reduction from white to red tiers vs. 40.0% and 43.3%).

**Figure 1.**
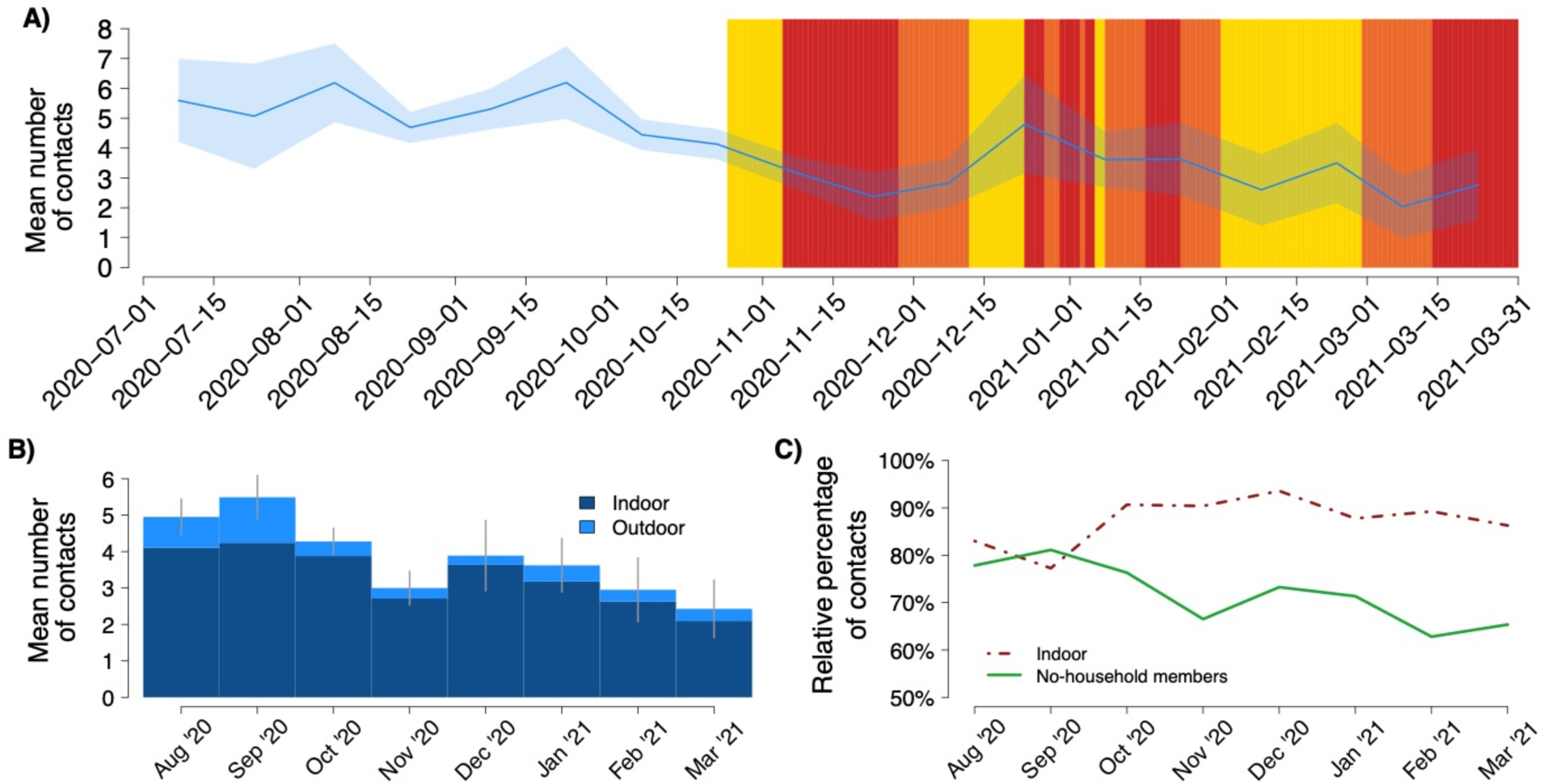
**A)** Colored rectangles represent the tiers assigned to Lombardy over the study period. The blue line represents the average number of daily contacts over time with respective 95% CI (shaded blue area). **B)** Bars represents the mean number of contacts reported by the study participants in different months, stratified by contacts occurred indoor and outdoor. Grey lines represent 95% confidence intervals of the overall number of daily contacts per person across different months. **C)** The dashed and solid lines represent respectively the proportions of contacts occurring indoor and of contacts occurring with no-household members across different months.

Individuals belonging to larger households reported a higher number of daily contacts in all tiers. While in the red tier, the mean number of reported contacts was just above the number of individuals cohabiting with the study participant.

Serological test results were mainly available for participants interviewed under the white tier, with those who tested negative for SARS-CoV-2 IgG antibodies prior to the interview reporting a slightly higher mean number of contacts (5.09 vs 4.10).

We found a progressive temporal decrease of both the mean daily number of contacts reported by study participants with no-household members and of the proportion of social encounters recorded indoor (see Figure 1). The observed dynamics is strongly related to changes characterizing the tiers assigned to Lombardy over the study period. However, our results also highlight that Christmas holidays might have temporally increased contacts occurring indoor with non-family members. While outdoor contacts accounted for about 15-20% of all contacts in late summer (August and September 2020), their contribution became less than 10% between October and December 2020.

To better disentangle the role of tiered restrictions in shaping the number of daily interactions, a negative binomial regression was applied to the number of contacts recorded by the study participants, adjusting for their sex, age, household size, employment status, and serological status. The same model was also separately applied to contacts recorded with household members and with non-household members. The resulting estimates (see Figure 2) suggest that, compared to what observed under the white tier, the daily number of contacts reported by the study participants decreases by 16.44% (95%CI: 5.37-26.13) under yellow tier, by 35.55% (95%CI: 24.21 -45.09) under the orange tier and by 45.60% (95%CI: 37.04-52.94) under the red tier (Figure 2). The mean number of contacts reported with household members did not significantly change across different tiers (reference: white tier; p-values of coefficients for yellow, orange, and red tiers: 0.40, 0.84 and 0.47, respectively). In contrast, the mean number of contacts with non-household members was found to significantly decrease respectively by 20.94 % (95%CI: 7.23 - 32.47), 44.03% (95%CI: 30.78 - 54.56) and 55.77% (46.48 - 63.36) under yellow, orange, and red tiers (Figure 2). Estimated model coefficients and relative confidence intervals are shown in Table S2 of the Appendix.

**Figure 2.**
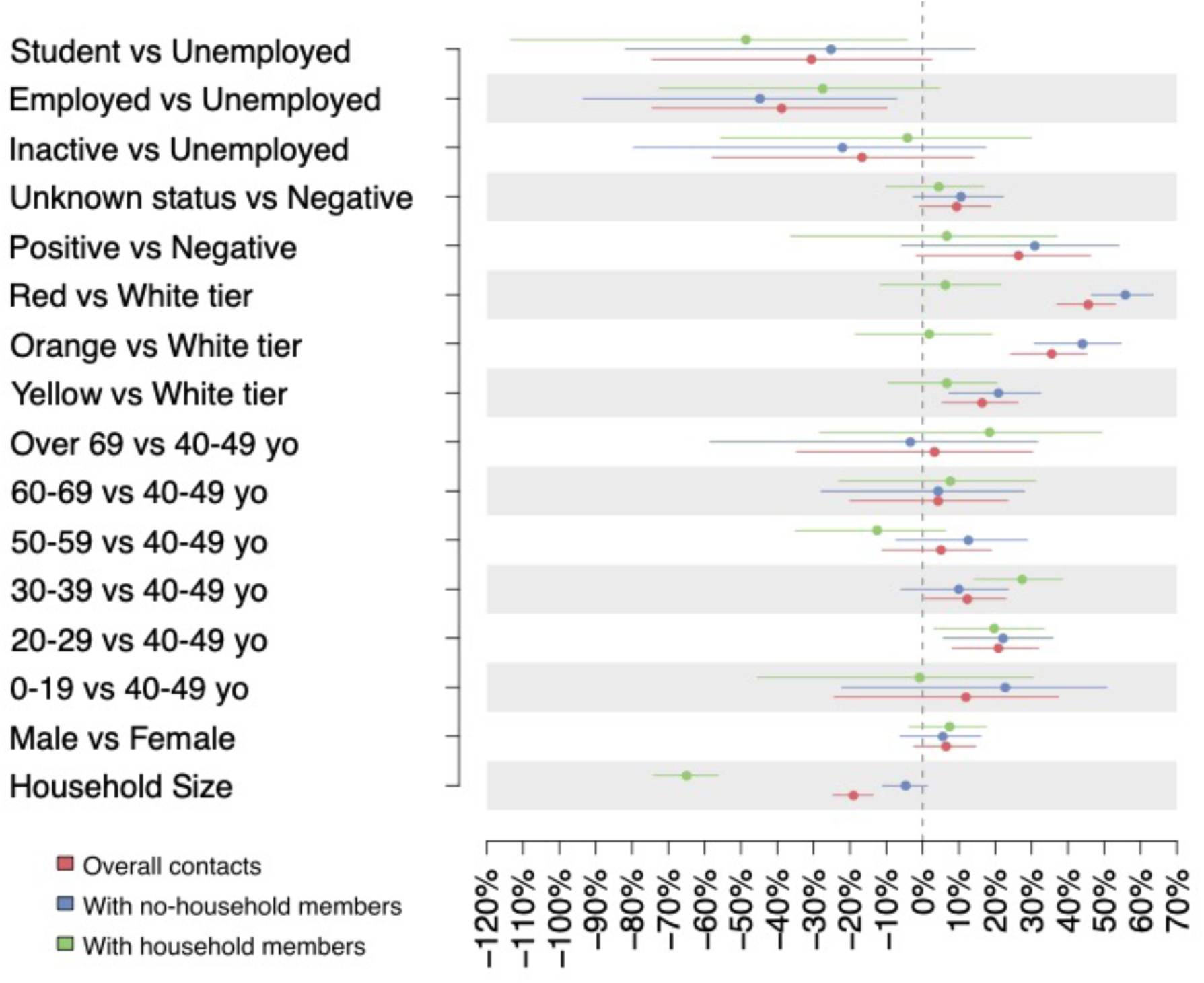
Percentage reduction of the daily number of contacts reported by study participants with respect to different covariates of interest, obtained as 1 minus the exponentiated coefficients estimated by a negative binomial regression applied to the overall number of contacts (red dots), the number of contacts occurred with no-household (blue dot) and with household members (green dots). Lines represent 95% confidence intervals.

Figure S2 in the Appendix show the results of a sensitivity analysis that considers the pre-tier period (October 26th - November 5th) separate from the yellow tier. The resulting estimates are in line with those obtained in the baseline analysis. A similar reduction in the average number of daily contacts was found for the yellow tier and the pre-tier period associated with the implementation of preventive restrictions in Lombardy. However, when these two periods are considered separately, the former shows a much broader variability around the estimated reduction.

### Age specific mixing patterns under different tiers

The analysis of contact patterns by age shows that higher restriction levels could markedly reduce both the number of intergenerational contacts had by the elderly (>60 years of age) and the intensity of assortative mixing in younger individuals. In children and adolescents (< 20 years of age), the latter phenomenon is likely related to the reinforcement of distance learning under more restrictive tiers. Beyond reducing the overall number of social interactions, higher restrictions levels were found to increase the relative contribution of contacts between young adults (aged 30-50 years) with individuals of similar age and with individuals younger than 10 years of age. This result is likely related to interactions occurring within the household between partners, and between parents and their children (Fig. 3A-D).

**Figure 3.**
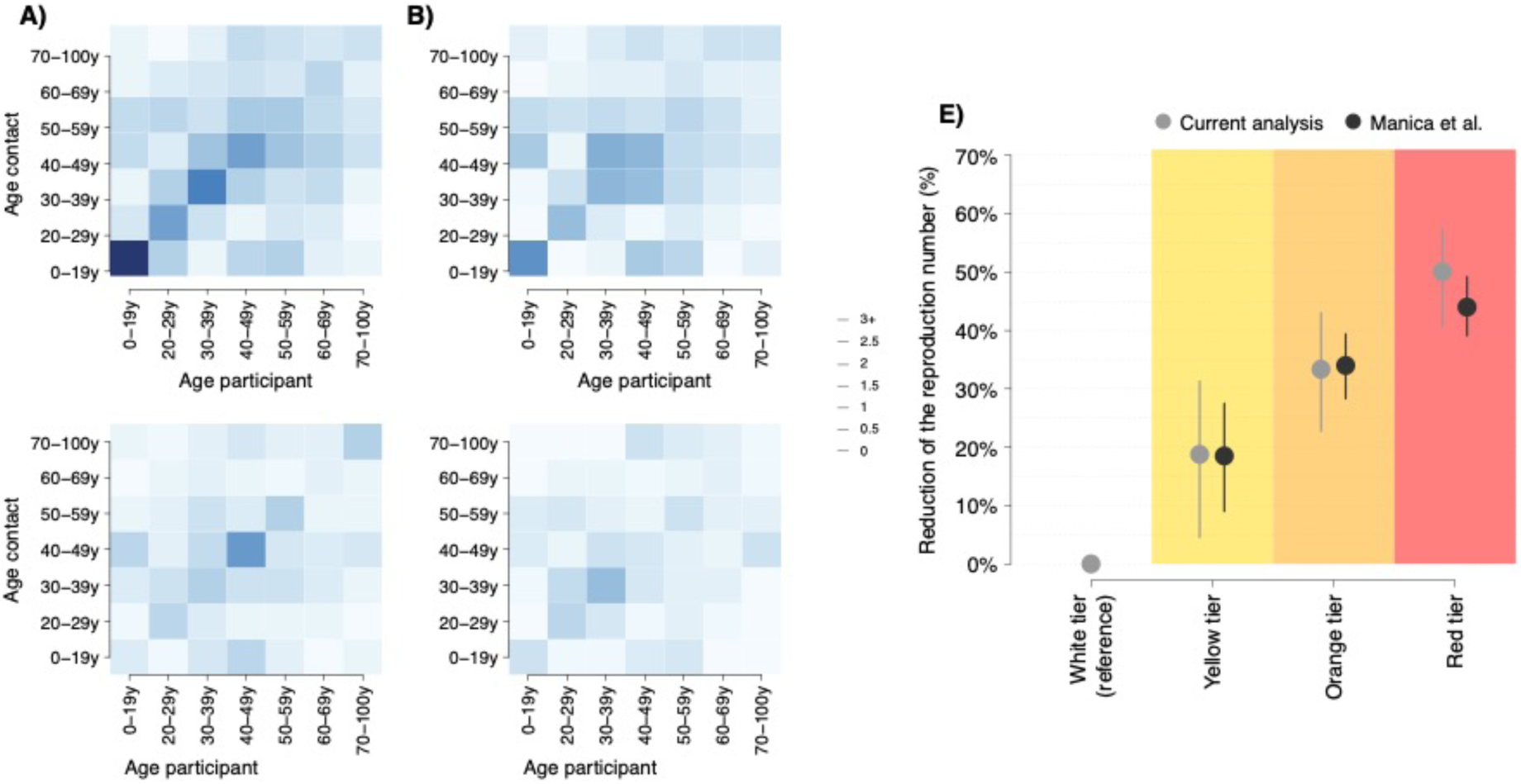
**A-D)** Average contact matrix representing the mean number of daily contacts reported by a participant in the age group *i* with individuals in the age group *j* under the white (A), the yellow (B), the orange (C) and the red tiers (D). **E)** Mean percentage reduction of SARS-CoV-2 reproduction number ascribable to the observed change of contact patterns (grey dots) and as estimated in [5] by analyzing the time series of the SARS-CoV-2 net reproduction number in Italy between October 30 and November 25, 2020 (black dots), under the yellow, the orange and the red tiers with respect to the white tier. Vertical lines represent 95% confidence intervals.

### Impact of restrictions on SARS-CoV-2 transmissibility

By comparing the transmission potential associated with contact patterns measured under different tiers, we found that - compared to what expected before the introduction of the tiered system – the SARS-CoV-2 reproduction number is expected to decrease by 18.7% (95%CI: 4.6-30.8), 33.4% (95%CI: 22.7-43.2), and 50.2% (95%CI: 40.9-57.7) under the yellow, orange, and red tiers, respectively (see Figure 3E). As shown in Figure S4 of the Appendix, these estimates are robust with respect to the assumption of homogeneous susceptibility at different ages, being respectively 19.3% (95%CI: 2.9-33.1), 35.9% (95%CI: 24.8-45.6), and 51.7% (95%CI: 41.5-59.4) under the yellow, orange, and red tiers.

Our estimates on the relative reduction of transmissibility potential are consistent with those obtained by a recent study that leverages epidemiological data from the Italian COVID-19 integrated surveillance system and mobility data to evaluate the impact of tiered restrictions on the net reproduction number [5]. According to this work, as shown in Figure 3, the relative reduction in the net reproduction number spans on average between 18.5% (95%CI: 9.0-27.5) under yellow tier restriction, 34.0% (95%CI: 28.3-39.4) under orange tier restrictions and 44.0% (95%CI: 39.1-49.2) under red tier restrictions.

The estimated contact matrices and reductions in the reproduction number of SARS-CoV-2 when considering separately the yellow tier and the pre-tier period associated with the implementation of preventive restrictions in Lombardy are in line with results obtained in the baseline analysis (see Figure S3 of the Appendix). Again, in this case, the estimate for the reduction in the reproduction number in the yellow tier shows much higher variability than that of the pre-tier period associated with preventive restrictions.

## DISCUSSION

The analyzed data was collected from a contact survey conducted in the metropolitan city of Milan between August 2020 and March 2021 through online interviews and encompasses all possible levels of restrictions coded by the tiered system introduced in Italy in November 2020. The collected data was used to provide estimates of the changes in the number of contacts and mixing patterns linked to the gradual tightening of non-pharmaceutical interventions and their potential impact on SARS-CoV-2 transmission. We found that the number of daily contacts per participant was significantly higher in late summer, when all social distancing measures except for the mandate of using PPE (personal protective equipment) in closed spaces were temporally relaxed in light of an improved epidemiological situation compared to spring 2020. Whether contact patterns follow a seasonal trend deserves further investigation.

Even in the period preceding the introduction of the three-tiered system, the average number of daily contacts shows a 4-fold reduction with respect to pre-pandemic levels estimated for Italy by the Polymod study [18]. According to the latter, the average number of daily contacts in Italy was 19.8 (SD=12.3), of which around 18% were school contacts. Age mixing patterns in the white tier are consistent with those estimated in the Polymod study [18], with marked assortative mixing among individuals under 30 years of age and intergeneration mixing between children and parents.

As tighter restrictions were implemented during the fall of 2020, a progressive reduction in the mean number of contacts recorded by study participants was observed: 16.44% under mild restrictions (yellow tier), 35.55% under moderate restrictions (orange tier) and 45.60% under strong restrictions (red tier). We estimated that the consequent impact on transmission might be quantified with a decrease of 18.7%, 33.4% and 50.2% in the SARS-CoV-2 reproduction number, respectively. A retrospective study analyzing SARS-CoV-2 epidemiological data at the level of Italian provinces has quantified the relative reduction of the net reproduction number associated to the adoption of different tiers between October and November 2020 [5]. Remarkably, the latter estimates well compare with what we have obtained in the present study, which analyzes contact patterns only (i.e., without any direct knowledge of the actual transmission patterns). First, this supports the relevance of mixing patterns as a predictor of infection transmission dynamics. Second, should these tiered restrictions be adopted again in the future and assuming a similar compliance of the population to the policy, our estimated change in mixing patterns could allow anticipating the impact of these interventions. Moreover, while inputting the estimated contact reduction in an epidemic model can provide insights on the impact of the implemented interventions foe any epidemiological situation (i.e., the model can simulate a certain level of vaccination, infection prevalence, etc.), the same may not be true for reductions of the reproduction numbers estimated under a specific epidemiological situation [5].

When interpreting our results, the following limitations should be considered. Firstly, the analyzed sample is affected by a strong selection bias. Indeed, study participants were enrolled among individuals who voluntarily registered to undergo a serological test. In addition, the conducted survey was restricted to the population of the metropolitan city of Milan. Consequently, our sample is not representative of the age distribution and of the household composition of the Italian population and results may not reflect social behavior adopted in less urbanized areas. In particular, crude estimates retrieved from our sample suggest that, in August 2020 (i.e., before introducing the tiered restriction system), the daily mean number of contacts was 74.5% lower than that estimated for Italy by the POLYMOD study in 2008 [18]. Although it is likely that, even in the absence of strong restrictions, current social interactions could strongly differ from those adopted in the pre-pandemic period [25], these numbers should be considered in light of the sampling procedure we adopted for data collection. For this reason, instead of relying on absolute numbers, our analysis focuses on investigating the relative changes in the contact patterns observed across different time periods and at different ages. To minimize potential biases, the potential impact of tier restrictions on the number of reported contacts was assessed by adopting a regression model, where a variety of potential confounding factors are considered.

An additional caveat should be also mentioned to correctly interpret the expected change of the SARS-CoV-2 reproduction number under different tiers. In fact, our estimates reflect only the impact of different restriction levels on the transmission when assuming constant epidemiological conditions over time. This means that we did not consider other factors which are known to strongly influence the spread of COVID-19, such as the level of immunity accrued over the course of the pandemic from natural infection or through the rollout of vaccination.

Despite the mentioned limitations, our study brings relevant contributions to the field. Our estimates could be used to approximate the expected contribution of different restriction levels in decreasing the transmission of SARS-CoV-2 independently from the current epidemiological situation, and they can therefore be instrumental for the control of future COVID-19 epidemics. Moreover, our approach could be generalizable to further monitor changes in social contacts during different epidemic phases or to evaluate alternative measures put in place to control the transmission of different infectious diseases.

## Data Availability

All data produced in the present study are available upon reasonable request to the authors

## ACKNOWLEDGMENTS

AM acknowledges support from the European Research Council (Consolidator Grant no. 101003183) and the Fondazione Invernizzi, Italy.

We thank all the staff at Centro Medico Sant’Agostino, and in particular Chiara Carrisi, Silvia Angeletti, Saverio Manin, Caterina Lurani, Marco Bertoncini, Antoine Sienczewski for their precious help in developing the platform and in the data collection process. We also thank the students Alice Rossini, Marta Pinzan, Luca Barbato, Antoine Sienczewski for their help in acquiring and processing the data.

