## Appendix for "Investigating the relationship between interventions, contact patterns, and SARS-CoV-2 transmissibility"

### 1. Restriction's timeline

Individual restrictions in place in the different tiers and in the pre-tier period between October 26th and November 5th, 2020, are shown in Table S1 below.

**Table S1.** List of the restrictions implemented in Lombardy during Covid-19 pandemic from June 2020 to April 2020. The table reports the difference of the restrictions in place in the four tires: the white tier, the pre- tier, the yellow tier, the orange tier and the red tier.

| | White tier | Pre-tiers (From 26th October to 5th November 2020) | Yellow tier | Orange tier | Red tier |
|---|---|---|---|---|---|
| **Face Masks** | Mandatory indoor spaces | Mandatory in/outdoor spaces | Mandatory in/outdoor spaces | Mandatory in/outdoor spaces | Mandatory in/outdoor spaces |
| **Individual Movements** | No restrictions | No restrictions | Stay-home mandate between 10 p.m. and 5 a.m. (except for work. health and other certified reasons) | Stay-home mandate between 10 p.m. and 5 a.m. and ban on movements between municipalities and to/from other regions (except for work. health and other certified reasons) | Full-day stay-home mandate and ban on movements between municipalities and to/from other regions (except for work. health and other certified reasons). |
| **Curfew** | None | None | From 10 p.m. to 5 a.m. | From 10 p.m. to 5 a.m. | All the day |
| **Retail and services** | Open | Shopping malls closed during weekends and | Shopping malls closed during weekends and | Shopping malls closed during weekends and holidays (with | All shops closed (with the exception of essential |

| | | | | | |
|---|---|---|---|---|---|
| | | holidays (with the exception of essential retail & services) | holidays (with the exception of essential retail & services) | the exception of essential retail & services) | retail & services) |
| **Schools & childcare** | Open | Distance learning in high schools | Distance learning in high schools and universities except when on-site attendance is essential (i.e.. for laboratory activities) | Distance learning in high schools and universities except when on-site attendance is essential (i.e.. for laboratory activities) | Distance learning in second and third grade of middle schools and in all grades of high schools and universities |
| **Bars serving food. Cafès & Restaurants** | Open | No service after 6 p.m. and take away allowed until 10 p.m. | No service after 6 p.m. and take away allowed until 10 p.m. | Closed. Take away allowed until 10 p.m. | Closed. Take away allowed until 10 p.m. |
| **Public transports** | 100% capacity for seats and between 25% and 50% reduction of the standing places (except school service) | 100% capacity for seats and between 25% and 50% reduction of the standing places (except school service) | 50% capacity reduction (except school service) | 50% capacity reduction (except school service) | 50% capacity reduction (except school service) |
| **Indoor recreational and cultural venues** | Museums and exhibitions open. Theaters | Museums and exhibitions open. Theaters | Closed | Closed | Closed |

| | | | | | |
|---|---|---|---|---|---|
| | and cinemas with limits of 1000 persons outdoor and 200 indoor | and cinemas closed | | | |
| **Gyms. pools & leisure venues** | Open | Gyms and pools closed. Other sports Centers open. | Closed except outdoor sport centers | Closed except outdoor sport centers | Individual outdoor training only (except sports events of national interest) |

#### 2. Demographic characteristics of the sample

In our sample, 0,22% of the study participants was between 0 and 10 years of age, 2.5% between 10 and 20 years of age, 22% between 20 and 30 years of age, 32.6% between 30 and 40 years of age, 19.5% between 40 and 50 years of age, 13.22% between 50 and 60 years of age, 6.73% between 60 and 70 years of age and, 3.13% older than 70 years.
Our sampling was slightly biased toward women, indeed 57.8% of the participants was women. Most of the participants were employed (77.4%), of the remaining part 8.2% was inactive, 10.1% consisted of students and 4,1% of unemployed individuals.
Finally, most of the participants have a household size of two (57.9%), the 19.4% have and household size of 3 and the 16.6% of 4, while only 2% of the sampled population lives alone and 4% lives in households larger than four people.

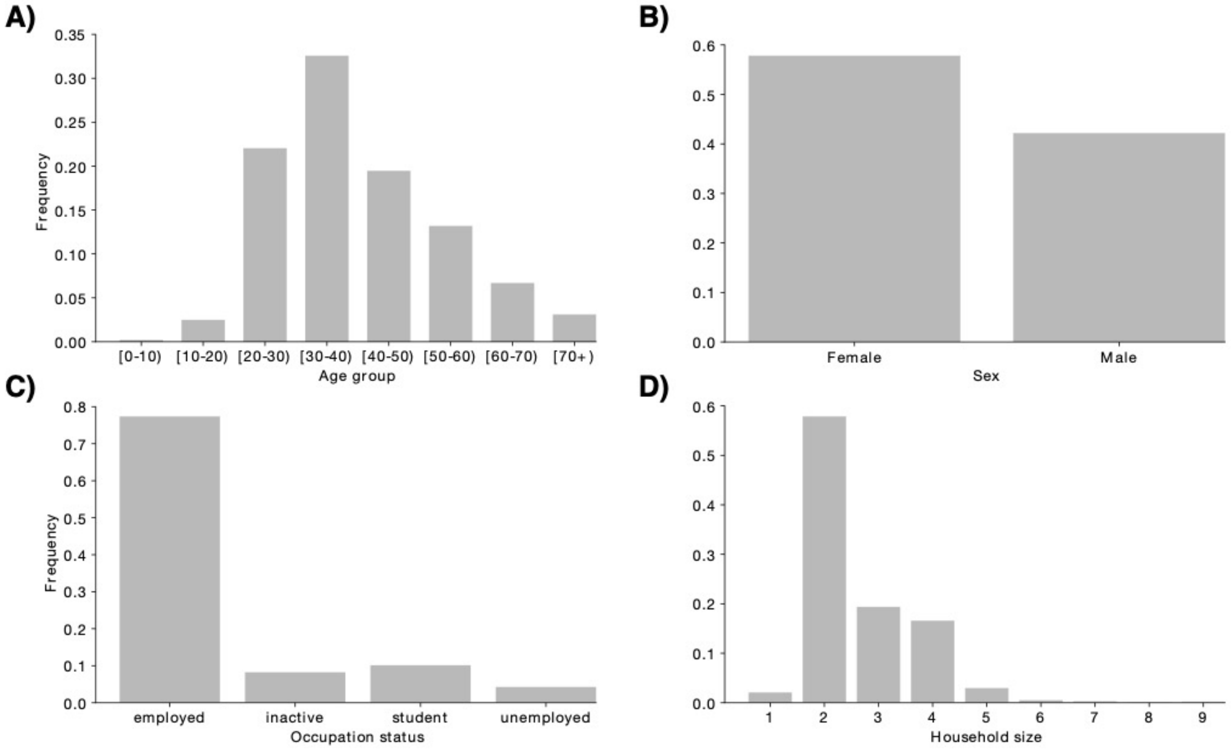

**Figure S1.** Demographic characteristics of the study participants. A) Age distribution B) Distribution of sex C) Distribution of occupation status, and (D) Distribution of household size.

#### 4. Adjustment of contact matrices for reciprocity

To robustly estimate the average number of observed contacts per person per day, we need to consider that the sample age distribution is different from age distribution of the population under study and to take into account the probability of an individual to be included in the sample. All the considered contact matrices were therefore adjusted for reciprocity, by applying the same approach used in [2] and detailed as follows.
Let $P_a$ denote the number of participants in the $a$-th age class and let $c_{a,\tilde{a}}(\mathrm{i})$ denote the number of contacts a specific study participant $i$ of age $a$ has with individuals of age $\tilde{a}$. The total number of contacts $T_{a,\tilde{a}}$ that all study participants of age $a$ have with individuals of age $\tilde{a}$

can be computed as

$$T_{a,\tilde{a}} = \sum_{\mathrm{i}=1}^{\mathrm{P}_a} c_{a,\tilde{a}}(\mathrm{i})$$

The average contacts an individual of age $a$ has with individuals of age $\tilde{a}$ can be approximated by the average contacts that a participant of age $a$ with individuals of age $\tilde{a}$ as follows:

$$C_{a,\tilde{a}} = \frac{T_{a,\tilde{a}}}{P_a}$$

In principle, $T_{a,\tilde{a}}$ can be different from $T_{\tilde{a},a}$. To correct matrices for symmetry we considered the probability of an individual to be included in the sample and we corrected the total number of contacts that all study participants of age $a$ have with individuals of age $\tilde{a}$ as a weighted average of the total contacts reported by participants of these two ages as follows:

$$T_{a,\tilde{a}}^{corrected} = \frac{\mathrm{P}_a \mathrm{N}_a T_{a,\tilde{a}} + \mathrm{P}_{\tilde{a}} \mathrm{N}_{\tilde{a}} T_{\tilde{a},a}}{\mathrm{P}_a + \mathrm{P}_{\tilde{a}}}$$

where $\mathrm{N}_a$ is the size of the age group $a$ in the population targeted by our contact survey. The adjusted average contacts an individual of age $a$ has with individuals of age $\tilde{a}$ was finally computed as

$$C_{a,\tilde{a}}^{corrected} = \frac{T_{a,\tilde{a}}^{corrected}}{\mathrm{N}_a}$$

#### 5. Uncertainty in contact matrices

To take into account sample variability, we computed 1,000 bootstrapped contact matrices. At each bootstrap iteration, we sampled with replacement 1063, 314, 178 and 230 interviews from those obtained in white, yellow, orange and red tiers respectively, choosing the age of the participant with probability proportional to the age distribution of the population residing in the metropolitan city of Milan. Then, we counted for each participant *i* of age group $a$ the number of contacts reported with individuals of age $\tilde{a}$ in the setting $x$, $c_{a,\tilde{a}}^{x}(i)$, and estimated the average number of contacts occurring in the setting $x$ between ages $a$ and $\tilde{a}$ from the following equation:

$$[\mathrm{Eq4}]\ C_{a,\tilde{a}}^{x} = \frac{\sum_{i=1}^{P_a} c_{a,\tilde{a}}^{x}(i)}{P_a}$$

where $P_a$ is the number of sampled participants of age group $a$.

#### 6. Estimates of the negative binomial regression models

Table S2 shows the estimated coefficients of the negative binomial regression on the number of overall contacts, the number of contacts with household and non-household members.

Table S2. Estimated coefficients, standard errors and p values as obtained by applying negative binomial regressions to the number of overall contacts, to the number of contacts with household members, and to the number of contacts with non-household members.

| | OVERALL | | | | CONTACTS WITH NON-HOUSEHOLD MEMBERS | | | | CONTACTS WITH HOUSEHOLD MEMBERS | | | |
|---|---|---|---|---|---|---|---|---|---|---|---|---|
| | Estimate | Std. Error | z | p value | Estimate | Std. Error | z | P value | Estimate | Std. Error | z | P value |
| Intercept | 1.030 | 0.146 | 7.039 | <0.001 | 1.128 | 0.188 | 5.985 | <0.001 | -1.477 | 0.186 | -7.952 | <0.001 |
| Household size | 0.174 | 0.023 | 7.604 | <0.001 | 0.045 | 0.030 | 1.492 | 0.136 | 0.500 | 0.024 | 21.070 | <0.001 |
| Male vs Female | -0.067 | 0.046 | -1.460 | 0.144 | -0.058 | 0.059 | -0.975 | 0.329 | -0.078 | 0.058 | -1.331 | 0.183 |
| 0-19 vs 40-49 | -0.127 | 0.174 | -0.731 | 0.465 | -0.259 | 0.231 | -1.122 | 0.262 | 0.008 | 0.187 | 0.041 | 0.967 |
| 20-29 vs 40-49 | -0.235 | 0.076 | -3.081 | 0.002 | -0.251 | 0.098 | -2.563 | 0.010 | -0.220 | 0.096 | -2.298 | 0.022 |
| 30-39 vs 40-49 | -0.133 | 0.065 | -2.054 | 0.040 | -0.105 | 0.083 | -1.270 | 0.204 | -0.320 | 0.084 | -3.809 | <0.001 |
| 50-59 vs 40-49 | -0.052 | 0.080 | -0.654 | 0.513 | -0.135 | 0.104 | -1.299 | 0.194 | 0.117 | 0.093 | 1.259 | 0.208 |
| 60-69 vs 40-49 | -0.044 | 0.116 | -0.378 | 0.705 | -0.044 | 0.149 | -0.294 | 0.769 | -0.079 | 0.148 | -0.535 | 0.593 |
| Over 69 vs 40-49 | -0.034 | 0.166 | -0.205 | 0.837 | 0.033 | 0.212 | 0.157 | 0.875 | -0.204 | 0.235 | -0.869 | 0.385 |
| Yellow vs White | -0.180 | 0.063 | -2.845 | 0.004 | -0.235 | 0.081 | -2.898 | 0.004 | -0.069 | 0.081 | -0.844 | 0.399 |
| Orange vs White | -0.439 | 0.081 | -5.391 | <0.001 | -0.580 | 0.105 | -5.503 | <0.001 | -0.019 | 0.097 | -0.197 | 0.844 |
| Red vs White | -0.609 | 0.075 | -8.143 | <0.001 | -0.816 | 0.097 | -8.401 | <0.001 | -0.065 | 0.090 | -0.717 | 0.474 |

| | | | | | | | | | | | | |
|---|---|---|---|---|---|---|---|---|---|---|---|---|
| Positive serology vs Negative | -0.307 | 0.162 | -1.894 | 0.058 | -0.371 | 0.209 | -1.771 | 0.077 | -0.069 | 0.195 | -0.355 | 0.723 |
| Unknown serological status vs Negative | -0.099 | 0.055 | -1.811 | 0.070 | -0.113 | 0.070 | -1.604 | 0.109 | -0.046 | 0.071 | -0.642 | 0.521 |
| Inactive vs Unemployed | 0.154 | 0.156 | 0.985 | 0.325 | 0.199 | 0.200 | 0.992 | 0.321 | 0.041 | 0.203 | 0.200 | 0.841 |
| Employed vs Unemployed | 0.328 | 0.117 | 2.793 | 0.005 | 0.370 | 0.151 | 2.455 | 0.014 | 0.243 | 0.152 | 1.590 | 0.112 |
| Student vs Unemployed | 0.267 | 0.149 | 1.794 | 0.073 | 0.224 | 0.192 | 1.166 | 0.243 | 0.396 | 0.184 | 2.148 | 0.032 |

### 7. Sensitivity analyses

#### 7.1. The pre-tier period associated with preventive restrictions in Lombardy

To better disentangle the role of pre-tier period associated with preventive restrictions in Lombardy from the yellow tier restrictions in shaping the number of daily interactions, a negative binomial regression was applied to the number of contacts experienced by the study participants, adjusting for their sex, age, household size, employment status and serological status, by considering the period between October 26th and November 5th, 2020, separately from the yellow tier. The same model was also applied to contacts recorded with household members and with non-household members.
The resulting estimates (see Figure S2) suggest that, compared to what observed under the white tier, the daily number of contacts reported by the study participants decreases by 19.22% (95%CI: 6.57-30.03) during the pre-tier period associated with preventive restrictions, by 10.54% (95%CI: -9.24-26.39) under yellow tier, by 35.54% (95%CI: 24.20 -45.08) under the orange tier and by 45.61% (95%CI: 37.05-52.95) under the red tier (Figure 2).

As expected, the mean number of contacts reported with household members did not significantly change across different tiers (reference: white tier; p-values of coefficients for pre-tier period associated with preventive restrictions and for yellow, orange, and red tiers: 0.41, 0.71, 0.84 and 0.47, respectively). In contrast, the mean number of contacts with non-household members was found to significantly decrease respectively by 23.87 % (95%CI: 8.21 - 36.61), 14.66 % (95%CI: -10.58-33.50), 44.01% (95%CI: 30.76 - 54.54) and 55.77% (46.49 - 63.36), respectively (Figure 2).

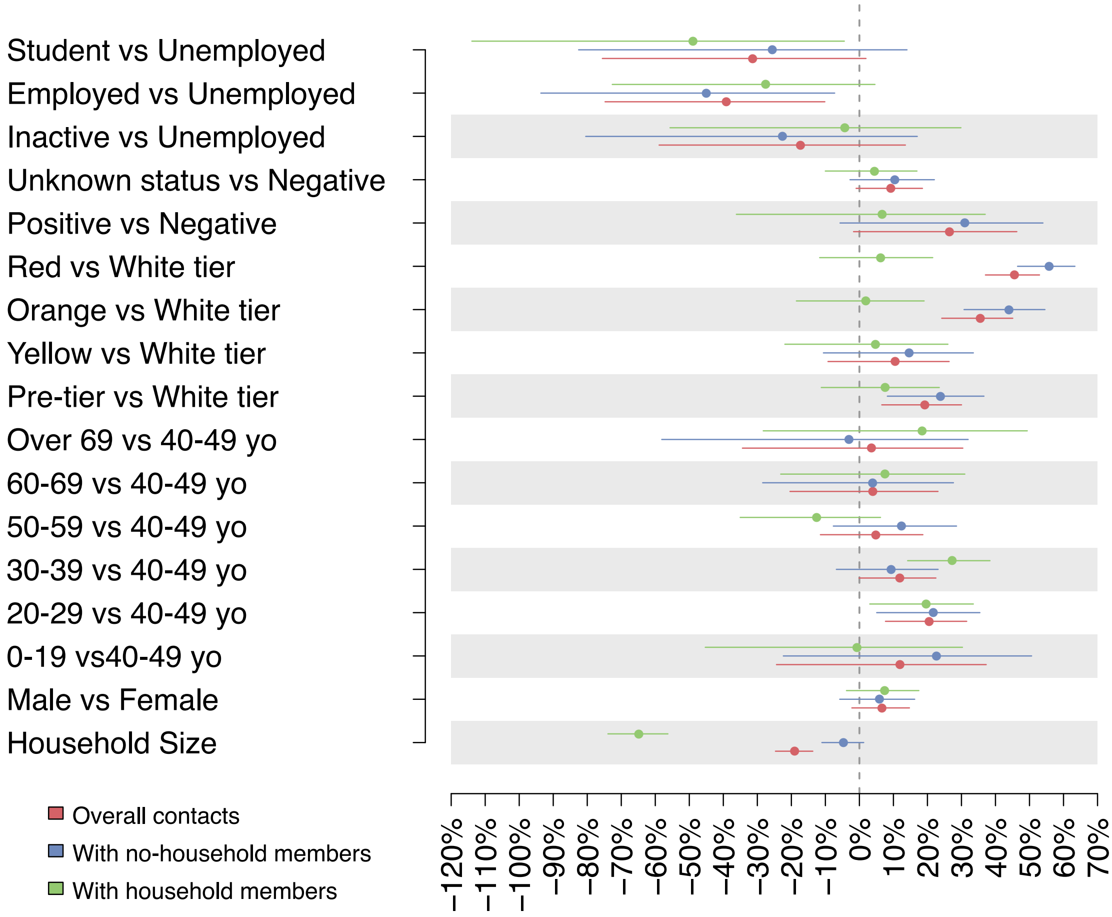

**Figure S2**. Percentage reduction of the daily number of contacts reported by study participants with respect to different covariates of interest, obtained as 1 minus the exponentiated coefficients estimated by a negative binomial regression applied to the overall number of contacts (red dots), the number of contacts occurred outside the household (blue dot) and within the household (green dots). Lines represent 95% confidence intervals.

By considering the same classification of periods, we estimated age-specific contacts matrices and the reduction in the SARS-CoV2 reproduction number under different levels of restrictions. Results are shown in Figure S3. The analysis of contact patterns by age clearly shows that higher restriction levels could markedly reduce both the number of intergenerational contacts experienced by the elderly (>60 years of age) and the intensity of assortative mixing in younger individuals. In children, the latter phenomenon is likely related to the reinforcement of distance learning under more restrictive tiers. Beyond reducing the overall number of social interactions, higher restrictions levels were found to increase the relative contribution of contacts between young adults (aged 30-50 years) with individuals of similar age and with individuals younger than 10 years of age.
Under the yellow tier, which in this sensitivity analysis coincide with the two weeks before Christmas break and February 2021, we did not estimate a large reduction in the number of contacts among different age strata of the population. Nonetheless it, is still observable from

Figure S3-C a reduction in overall intergeneration mixing and an increase in the relative contribution of contacts between young adults (aged 30-50 years) with individuals of similar age and with individuals younger than 10 years of age.

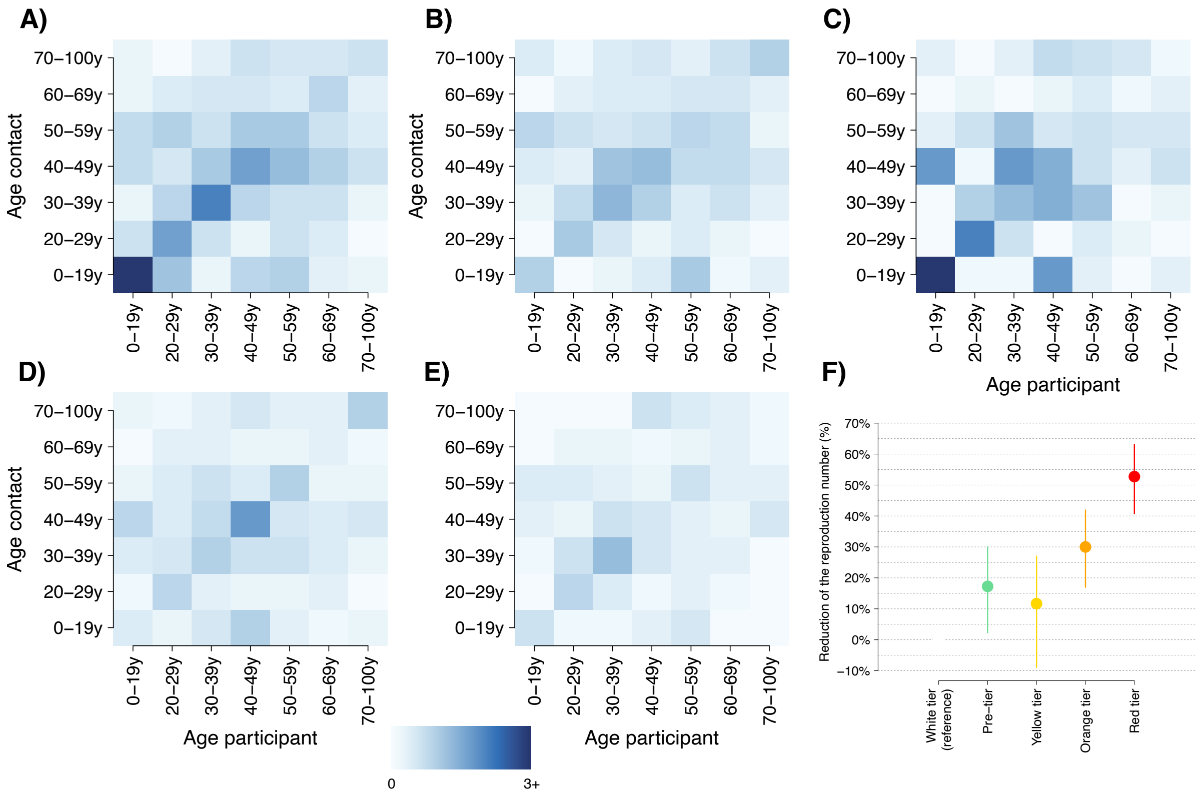

**Figure S3.** Average contact matrix representing the mean number of daily contacts reported by a participant in the age group i with individuals in the age group j under the white (A), the pre-tier period associated with preventive restrictions (B) the yellow (C), the orange (D) and the red tiers (E). Average percentage reduction of the SARS-CoV-2 reproduction number ascribable to the observed change of contact patterns under the pre-tier (green) period associated with preventive restrictions, the yellow, the orange and the red tiers with respect to the white tier. Lines represent 95% bootstrapped confidence intervals (F).

By comparing the transmission potential associated with contact patterns measured under different tiers, we found that - compared to what expected before the introduction of tighter restrictions in the Lombardy region – the SARS-CoV-2 reproduction number is expected to decrease by 17.2% (95%CI: 3.8-29.4), 11.4% (95%CI: -9.1-28.8), 30.2% (95%CI: 16.9-41.4), and 52.4% (95%CI: 40.0-62.8) under the pre-tier period associated with preventive restrictions, the yellow tier, the orange tier and red tier, respectively (see Figure S3).

#### 7.2 Homogeneous susceptibility

By comparing the transmission potential associated with contact patterns measured under different tiers, under the assumption of homogeneous susceptibility, we found that - compared to the white tier – the SARS-CoV-2 reproduction number is expected to decrease by 19.3% (95%CI: 2.9-33.1), 35.9% (95%CI: 24.8-45.6), and 51.7% (95%CI: 41.5-59.4) under the yellow, orange and red tiers, respectively (see Figure S4).

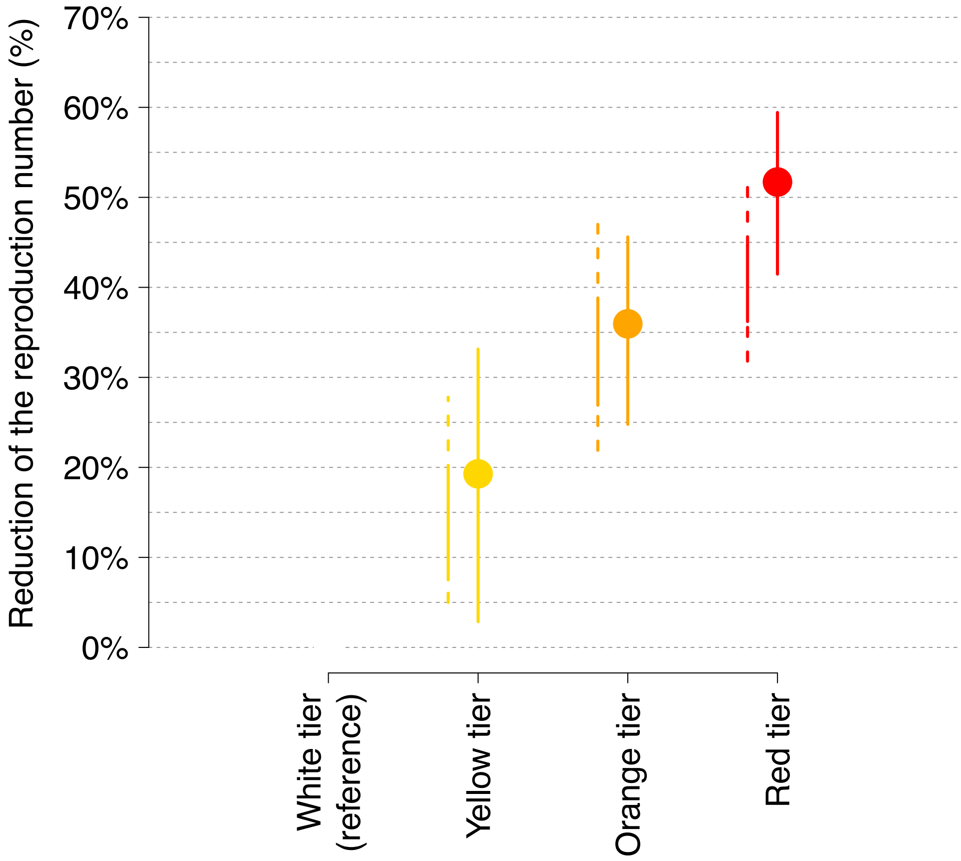

**Figure S4.** Average percentage reduction of the SARS-CoV-2 reproduction number ascribable to the observed change of contact patterns under the yellow, the orange and the red tiers with respect to the white tier when assuming homogeneous susceptibility across different ages. Lines represent 95% bootstrapped confidence intervals.

[1] Diekmann O, Heesterbeek JA, Metz JA. On the definition and the computation of the basic reproduction ratio R0 in models for infectious diseases in heterogeneous populations. J Math Biol. 1990;28(4):365–82.

[2] Melegaro A, Del Fava E, Poletti P, Merler S, Nyamukapa C, Williams J, Gregson S, Manfredi P. Social Contact Structures and Time Use Patterns in the Manicaland Province of Zimbabwe. PLoS One. 2017 Jan 18;12(1):e0170459.
